# SARS Coronavirus-2 microneutralisation and commercial serological assays correlated closely for some but not all enzyme immunoassays

**DOI:** 10.1101/2020.12.07.20245696

**Authors:** Gregory J Walker, Zin Naing, Alberto Ospina Stella, Malinna Yeang, Joanna Caguicla, Vidiya Ramachandran, Sonia R Isaacs, David Agapiou, Rowena A Bull, Sacha Stelzer-Braid, James Daly, Iain B Gosbell, Veronica C Hoad, David O Irving, Joanne M Pink, Stuart Turville, Anthony D Kelleher, William D Rawlinson

## Abstract

**Background:** Serological testing for SARS-CoV-2 specific antibodies provides important research and diagnostic information relating to COVID-19 prevalence, incidence, and host immune response. A greater understanding of the relationship between functionally neutralising antibodies detected using microneutralisation assays and binding antibodies detected using scalable enzyme immunoassays (EIA) is needed in order to address protective immunity post-infection or vaccination, and assess EIA suitability as a surrogate test for screening of convalescent plasma donors. We assessed whether neutralising antibody titres correlated with signal cut-off ratios in five commercially available EIAs, and one in-house assay based on expressed spike protein targets.

**Methods:** Sera from individuals recovered from patients or convalescent plasma donors who reported laboratory-confirmed SARS-CoV-2 infection (n=200), and negative control sera collected prior to the COVID-19 pandemic (n=100) were assessed in parallel. Performance was assessed by calculating EIA sensitivity and specificity with reference to microneutralisation.

**Results:** Neutralising antibodies were detected in 166 (83%) samples. Compared with this, the most sensitive EIAs were the Cobas Elecsys Anti-SARS-CoV-2 (98%) and Vitros Immunodiagnostic Anti-SARS-CoV-2 (100%), which detect total antibody targeting the N and S1 antigens, respectively. The assay with the best quantitative relationship with microneutralisation was the Euroimmun IgG.

**Conclusions:** These results suggest the marker used (total Ab vs IgG vs IgA), and the target antigen are important determinants of assay performance. The strong correlation between microneutralisation and some commercially available assays demonstrate their potential for clinical and research use in assessing protection following infection or vaccination, and use as a surrogate test to assess donor suitability for convalescent plasma donation.

## Introduction

The easing of COVID-19 control measures requires extensive surveillance for the early detection of new clusters, as well as an understanding of the level and duration of protective immunity in the community. Serological testing for SARS-CoV-2 specific antibodies is an important tool that serves multiple diagnostic and research purposes including: i) confirmation of suspected infection, ii) informing public health policy by determining the true infection rate (symptomatic and asymptomatic cases) occurring within a population, iii) assessing seroconversion following infection or vaccination and iv) as a potential scalable screening test to determine suitablitiy for convalescent plasma donation (1-4).

It is important to decipher the neutralising capability of developed SARS-CoV-2 specific antibodies to understand whether the host response will provide sufficient protection from future reinfection. Neutralising antibodies can be detected using the microneutralisation assay and plaque reduction neutralisation test. These assess the ability of patient-derived serum samples containing SARS-CoV2 specific antibody to inhibit infection of cells cultured in vitro (5). These virus neutralisation tests require the handling of replication-competent SARS-CoV-2 in specialized containment laboratories (biosafety level 3, at minimum (6)), and for this reason are impractical to scale. Commercially available serology tests such as enzyme immunoassays (EIA) are faster and less laborious than traditional culture-based methods, which is advantageous in the diagnostic laboratory setting (7). However, these assays do not differentiate between binding antibodies and neutralising antibodies (8). The detection of binding antibodies does not necessarily confer virus-neutralisation or protection against virus replication in the infected host, and traditional virus neutralisation tests remain the reference standard (9). Correlation of binding antibodies detected using EIA with neutralising antibody titres will be crucial for population-level screening of seroconversion and assessment of herd-immunity following vaccination and rapid assessment of the suitablity of convalescent plasma donors (4, 10).

In response to demand for SARS-CoV-2 serological testing kits, numerous assays have been released under relaxed regulatory assessment criteria (1). Validation studies by end-users are important to assess the performance characteristics of these new commercial assays, and to determine the correlation between EIAs and neutralising antibody titres. To date, a small number of studies have validated a range of commercially available SARS-CoV-2 serological assays against a live-virus neutralisation test (11-15). Assays assessed in these publications incorporate automated platforms as well as serology-based point of care tests and have some, but limited crossover with the comparison of EIAs in the present study. Others tested too few samples to effectively correlate EIA resuts with neutralising antibody titres (16-20), lacked comparison of head-to-head EIAs (21, 22), or used live-virus neutralisation as a reference standard primarily to validate assays developed in-house (23-26). Here we report on the performance of a unique set of five commercially available SARS-CoV-2 serological assays and an in-house developed EIA, with reference to a reference-standard microneutralisation assay.

## Methods

### Sample collection and testing

All samples were received by the Serology and Virology Division at the Prince of Wales Hospital Randwick, Australia. Two hundred sera were collected from laboratory-confirmed COVID-19 patients (n=157) between March and June of 2020. The majority were recruited as convalescent plasma donors (self-reported laboratory confirmed infection) and tested as part of the release test to ensure donor suitablity on behalf of Australian Red Cross Lifeblood (161 samples from 124 donors). Donor samples were collected 37 to 101 days post-test postivitity date (mean=60·5). The donors ranged in age from 20 to 78 years old (mean =45·3 years) with 54.6% being males. A smaller proportion of serum (39/200), were obtained from COVID-19 patients 1 to 47 days post-laboratory-confirmed diagnosis. An additional 100 sera were obtained from patients prior to the COVID-19 pandemic between 2016 and 2018 (control cohort). This included 25/100 samples serologically positive for antibodies to common respiratory viruses (Supplementary Table 1). Antibodies to SARS-CoV-2 in samples were measured using a microneutralisation assay at two dilutions (1:40 and 1:80), and up to six other immunoassays including an in-house delveloped EIA. Commercially available assays were performed according to the manufacturer’s instructions, using kits of the same lot number for all assays. Samples returning equivocal/borderline results (as per the manufacturer-specified range) were not included in sensitivity and specificity calculations.

**Table 1.** Performance of commercially available SARS-CoV-2 serological assays

|  | Microneutralisation | Cobas total Ab | Vitros total Ab | Abbott IgG | In-house IgG | Euroimmun IgG | Euroimmun IgA |
| --- | --- | --- | --- | --- | --- | --- | --- |
| **Platform** | Cell culture | ECLIA | CLIA | CMIA | ELISA | ELISA | ELISA |
| **Antigen** | - | N | S1 | N | RBD | S1 | S1 |
| **All COVID-19 samples** |  |  |  |  |  |  |  |
| Total | 200 | 197 | 98 | 199 | 94 | 132 | 96 |
| Positive | 166 | 175 | 88 | 139 | 79 | 106 | 51 |
| Negative | 34 | 22 | 10 | 60 | 11 | 24 | 32 |
| Equivocal | - | - | - | - | 4 | 2 | 13 |
| Positivity (%) | 83.0 | 88.8 | 89.8 | 69.8 | 87.8 | 81.5 | 61.4 |
| Sensitivity <sup>1</sup> (%) | - | 98.2 | 100.0 | 78.8 | 96.1 | 94.3 | 69.4 |
| **Convalescent samples <sup>2</sup>** |  |  |  |  |  |  |  |
| Total | 167 | 167 | 73 | 167 | 74 | 109 | 73 |
| Positive | 151 | 160 | 72 | 123 | 65 | 93 | 40 |
| Negative | 16 | 70 | 1 | 43 | 6 | 66 | 24 |
| Equivocal | - | - | - | - | 3 | 2 | 9 |
| Positivity (%) | 90.4 | 95.8 | 98.6 | 73.7 | 91.5 | 86.9 | 62.5 |
| Sensitivity <sup>1</sup> (%) | - | 98.7 | 100.0 | 76.8 | 95.3 | 93.6 | 65.0 |
| **Negative samples** |  |  |  |  |  |  |  |
| Total | 100 | 100 | 99 | 100 | 100 | 100 | 100 |
| Positive | 0 | 0 | 0 | 0 | 4 | 0 | 6 |
| Negative | 100 | 100 | 99 | 100 | 96 | 100 | 92 |
| Equivocal | - | - | - | - | - | - | 2 |
| Specificity <sup>1</sup> (%) | 100.0 | 100.0 | 100.0 | 100.0 | 96.0 | 100.0 | 93.9 |
| **PPV (%)** | - | 93.7 | 90.9 | 93.5 | 89.2 | 94.3 | 87.7 |
| **NPV (%)** | - | 97.5 | 100.0 | 78.1 | 97.2 | 95.1 | 77.3 |

<sup>1</sup> Sensitivity and specificity calculated against reference-standard virus neutralisation test
<sup>2</sup> Samples were considered convalescent if they were collected $\geq 14$ days post-NAT detection
Equivocal results were excluded from sensitivity and specificity calculations
ECLIA, electrochemiluminescence immunoassay; CLIA, chemiluminescent immunoassay; CMIA, chemiluminescent microparticle immunoassay; ELISA, enzyme linked immunosorbent assay; N, nucleoprotein; S1, spike glycoprotein subunit 1; RBD, receptor binding domain; PPV, positive predictive value; NPV, negative predictive value

### Virus microneutralisation assay

Dilutions of test serum were prepared on a 96-well plate in duplicate in viral culture media (MEM + 2% fetal bovine serum + 1x penicillin-streptomycin-glutamine). The dilutions were incubated for one hour at 37°C with an equal volume of 200 TCID_50_ SARS-CoV-2 isolate. A suspension of Vero E6 cells containing 2×10^4^ cells was added to each well, and plates were incubated at 37°C (5% CO2) for three days. The plates were observed for cytopathic effect and the neutralisation titre determined as the dilution that conferred complete protection from infection in both replicates. Neutralising titres of 1:40 and above were considered positive.

### Cobas Elecsys Anti-SARS-CoV-2

The Elecsys Anti-SARS-CoV-2 (Roche Diagnostics, Australia) is an electrochemiluminescence immunoassay (ECLIA) for the detection of total antibody against the N protein of SARS-CoV-2 in serum and plasma. The platform provided a readout indicating whether the sample measurement is above or below the signal cutoff, and was interpreted as positive (≥1·0) or negative (<1·0).

### Vitros Immunodiagnostic Anti-SARS-CoV-2

The Vitros Immunodiagnostic Anti-SARS-CoV-2 (Ortho-Clinical Diagnostics, Australia) is a chemiluminescent immunoassay (CLIA) utilizing a recombinant SARS-CoV-2 S1 protein to measure total antibody present in serum and plasma. The platform provided a readout indicating whether the sample measurement was above or below the signal cutoff, and was interpreted as positive (≥1·0) or negative (<1·0).

### Abbott Architect SARS-CoV-2 IgG

The Architect SARS-CoV-2 IgG (Abbott Diagnostics, Australia) is a chemiluminecent microparticle immunoassay (CMIA) for the detection of IgG antibodies to the nucleocapsid (N) protein of SARS-CoV-2 in serum and plasma. The platform calculated a result by dividing the chemiluminescent signal from each sample with a calibrated signal. The unit for the assay is Index (S/C) and the result was interpreted as positive (≥1·4) or negative (<1·4).

### Euroimmun Anti-SARS-CoV-2 ELISA

The Anti-SARS-CoV-2 (Euroimmun, Germany) is an enzyme linked immunosorbent assay (ELISA) based platform that utilizes recombinant S1 protein to bind anti-SARS-CoV-2 antibodies in serum or plasma. Separate kits for the detection of IgG and IgA were used. Photometric measurement of colour intensity was used to calculate a ratio of the sample over the calibrator. The ratio was interpreted as positive (≥1·1), borderline (≥0·8 -<1·1), or negative (<0·8).

### In-house RBD assay

The in-house ELISA was performed by coating 96-well microtiter ELISA plates with biotinylated RBD antigen (Supplementary information), which bound SARS-CoV-2 specific antibodies. Diluted serum (1:101) or controls were added to respective wells for one hour. Wells were aspirated and washed three times with wash solution (PBS + 1% Tween 20). A secondary antibody (antihuman-IgG conjugated with the enzyme alkaline phosphatase, Virion/Serion) was then added to wells for 30 minutes to detect and bind the immune complex. The wash step was then repeated, before the addition of substrate solution (p-nitrophenylphosphate, Virion/Serion) for 30 minutes. The stopping solution was then added (<0·1 N sodium hydroxide and 40 mM EDTA, Virion/Serion) and absorbance was read at 405/620 nm. The cutoff optical density (OD) was set at 0·2 above the negative control OD. Samples were considered positive if the absorbance value was equal to or higher than the cutoff value.

### Statistical Analyses

The performance of the commercial assays were compared using results of the microneutralisation assay as the reference-standard. For sensitivity, only microneutralisation-positive samples were used in calculations. Assay specificity and cross-reactivity were assessed using samples from the negative-control cohort. Equivocal results were excluded from sensitivty and specificity calculations. Figures including optical density ratios and ROC curves were generated in GraphPad Prism 9. For the purpose of this analysis, serum collected ≥14 days post-laboratory confirmed COVID-19 diagnosis was considered convalescent.

Ethics approval to compare and validate antibody SARS-CoV-2 assays as part of a larger project to collect, manufacture and supply convalescent plasma to patients enrolled in clinical trials and for COVID-19 Immunoglobulin was approved by the Lifeblood Ethics Committee (approval number Hoad 30042020).

## Results

Neutralising antibodies were detected in 166/200 (83%) COVID-19 confirmed sera (Table 1). There were 151/167 (90%) convalescent sera (≥14 days) positive for neutralising antibodies including 112 (67%) which were neutralising at the highest dilution tested (1:80). No samples in the control cohort were positive for neutralising antibodies.

Across all assays SARS-CoV-2 antibodies were detected in 182/200 (91%) samples tested, with individiual EIA positivity ranging between 61-90% (Table 1). There were five convalescent sera from NAT-confirmed COVID-19 patients that returned negative results by microneutralisation and all EIAs tested (samples tested on ≥3 EIAs).

The sensitivity values of the commercially available assays against the microneutralisation reference-standard ranged from 69-100%, with assays measuring total antibody being most sensitive (Table 1). There was little difference in the sensitivity of assays in detecting neutralising antibody between acute and convalescent COVID-19 samples. Optical density ratios for each EIA, at varying neutralising antibody titres are shown in Figure 1. The Euroimmun IgG assay had the highest positive predictive value (94%) and displayed the best quantitative relationship with microneutralisation (Figure 2). All EIAs other than the Euroimmun IgA (94%) and in-house ELISA (96%) displayed 100% specificity in testing the negative control cohort.

**Figure 1.**
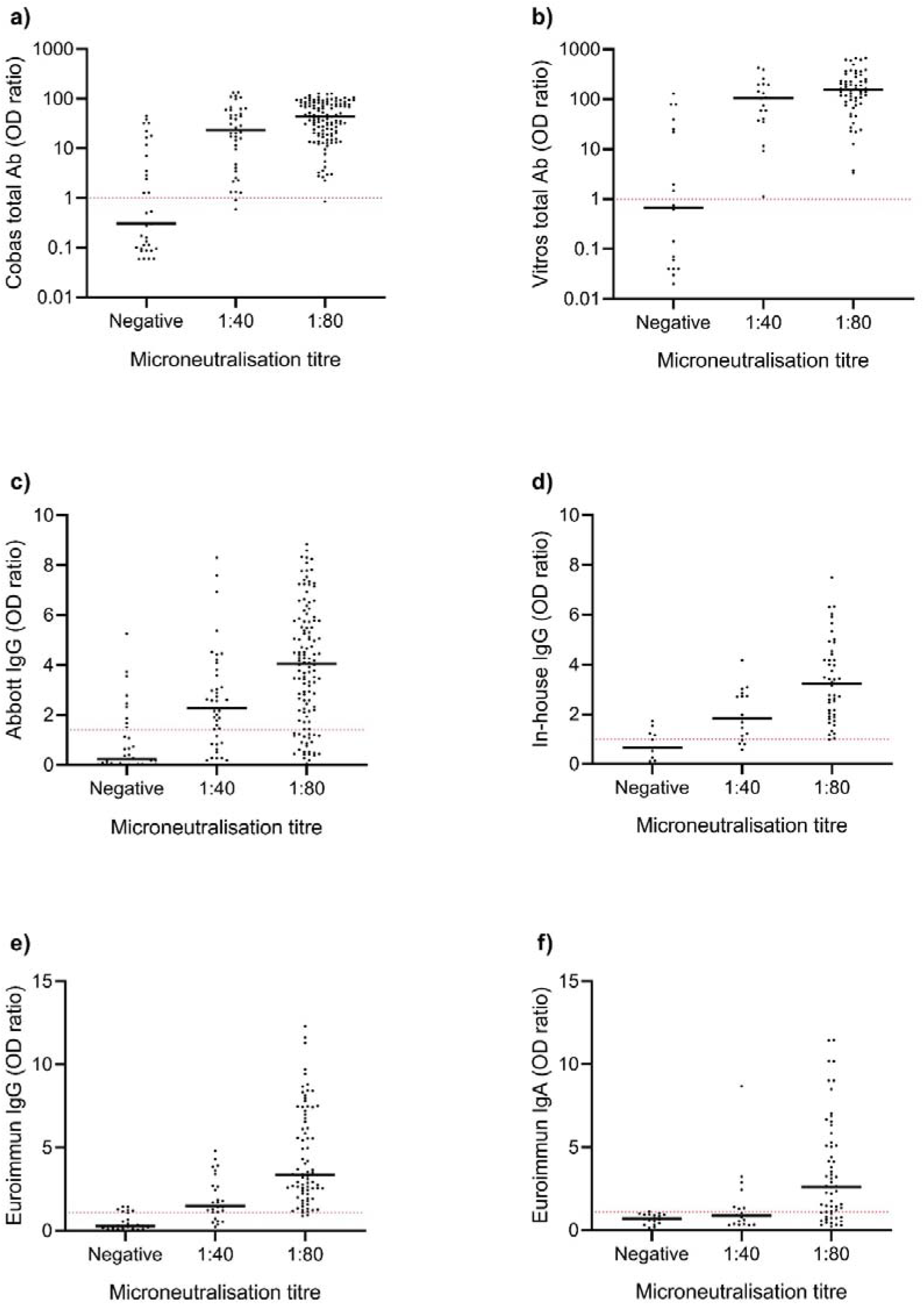
Optical density ratios of SARS-CoV-2 enzyme immunoassays (EIA) at varying neutralising antibody titres. Serum from COVID-19 confirmed patients were assayed using microneutralisation and (A) Cobas Elecsys Anti-SARS-CoV-2, (B) Vitros Immunodiagnostic Anti-SARS-CoV-2, (C) Abbott Architect SARS-CoV-2 IgG, (D) an In-house IgG ELISA, (E) Euroimmun Anti-SARS-CoV-2 IgG ELISA, and (F) Euroimmun Anti-SARS-CoV-2 IgA ELISA. Dashed line indicates the positive cut-off value of each assay determined by the manufacturer. The width of the scatter plot is proportionate to the number of data points at a given value, and the median EIA optical density shown. OD, optical density.

**Figure 2.**
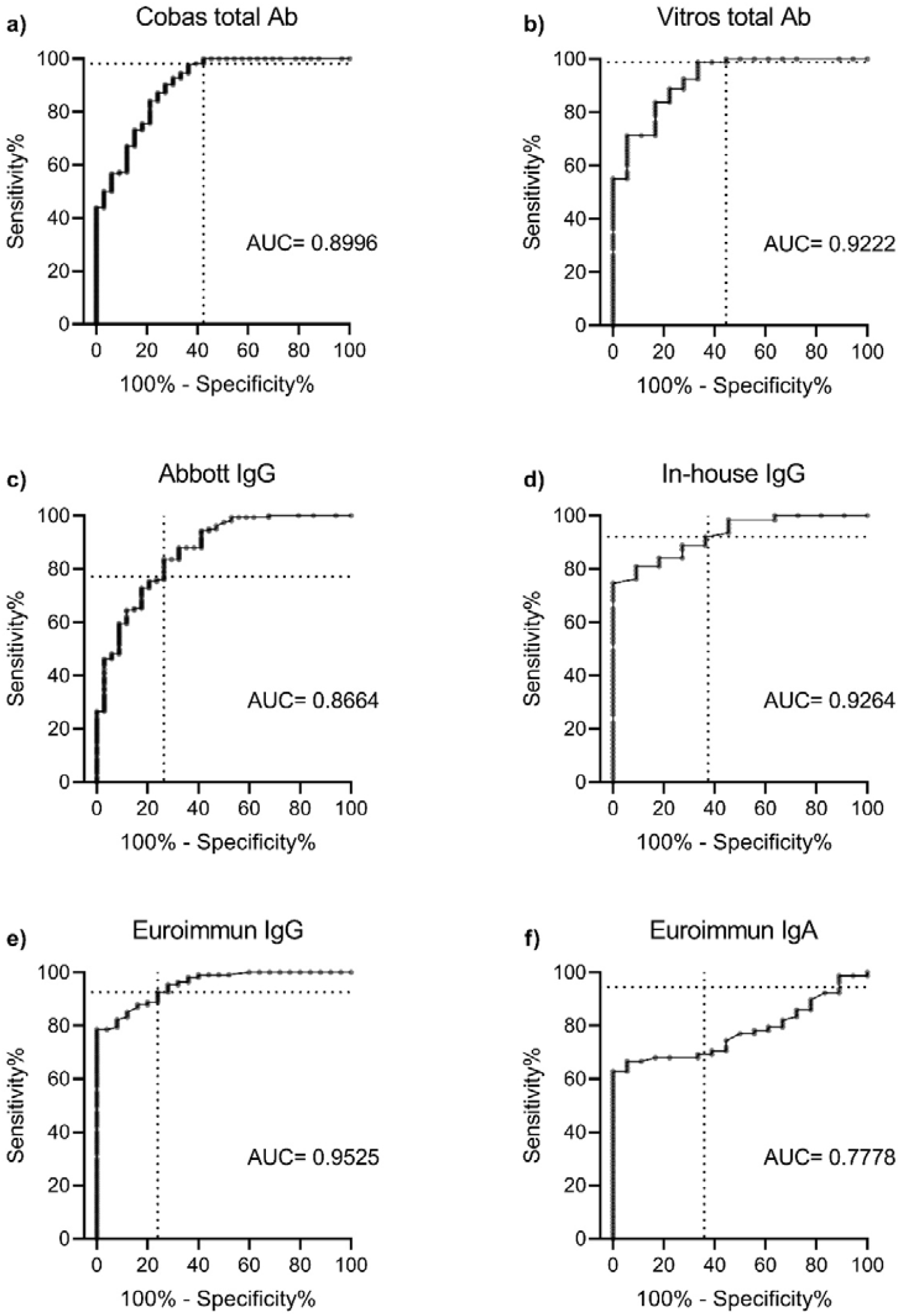
ROC curves for each enzyme immunoassay (A) Cobas Elecsys Anti-SARS-CoV-2, (B) Vitros Immunodiagnostic Anti-SARS-CoV-2, (C) Abbott Architect SARS-CoV-2 IgG, (D) an In-house IgG ELISA, (E) Euroimmun Anti-SARS-CoV-2 IgG ELISA, and (F) Euroimmun Anti-SARS-CoV-2 IgA ELISA. Note: Data are presented for up to 200 COVID-19 confirmed sera using microneutralisation as the reference-standard. Euroimmun IgG had the best correlation with the detection of neutralising antibody (AUC= 0·9525). Dashed line represents assay performance at the positive cut-off value specified by the manufacturer. ROC, receiver operating characteristic; AUC, area under curve.

## Discussion

Serological assays need to have validated performance characteristics in assessing SARS CoV-2 infection for diagnosis, surveillance and correlation with other assays. Given neutralisation activity is important for immunity and for potential therapeutic options such as convalescent plasma and immunoglobulin production, the major strength of our study is that it is one of the few to validate serological assays for SARS-CoV-2 against a virus neutralisation test. On reference testing, neutralising antibodies were detected in 83% of samples, compared to EIA which detected SARS-CoV-2 antibodies in 91% of samples across all assays. This was unsurprising considering that not all patients with COVID-19 develop neutralising antibodies to infection (27). For this reason, all assays produced false positives in reference to microneutralisation, with Euroimmun IgG displaying the highest positive predictive value and best correlation with neutralising antibody titres. Clinically, EIAs that correlate strongly with microneutralisation are of importance considering that the detection of neutralising antibodies may be useful in informing return to work and the discontinuation of transmission-based precautions (1).

For the aim of detecting protective neutralising antibodies, the S1-based Vitros Immunodiagnostic and the N-based Cobas Elecsys assays were most sensitive. A previous study suggested that assays targetting RBD and N proteins were more sensitive and better correlated with neutralisation titres than those targeting the S1 protein (23). Our testing of 300 samples found assay sensitivity to be dependent on assay marker (total Ab vs IgG vs IgA), and quantitative relationship dependent on the target antigen used – with the three spike-based assays best correlating with neutralising antibody titre (Figure 2).

In line with other validation studies using composite reference standards (28-30), the Euroimmun anti-SARS-CoV-2 IgG ELISA was highly sensitive and specific. However, the reported performance of the Euroimmun anti-SARS-CoV-2 IgA ELISA is mixed, and here we found it to have low sensitivity. IgG response is longer lived (31) and these findings may be reflective of our cohort, which was primarily made up of convalescent serum collected more than 36 days following laboratory-confirmed infection. The performance of the Architect IgG assay in detecting neutralising antibody was moderate, and in convalescent samples positivity was well below the sensitivity values advertised by the manufacturer (74% vs 100%). This highlights the importance of independent validation studies comparing a range of assays against multiple reference standards. Interestingly, there were five convalescent sera from reported laboratory-confirmed COVID-19 patients that returned negative results by microneutralisation and all EIAs tested. This also occurred in a validation study and could be explained by a failure to develop a measureable systemic antibody response, clearance of infection via other immune mechanisms, or an initial false-positive RT-qPCR result (32). Whilst donors in our study were required to confirm they had laboratory confirmed infection to donate, proof in terms of visualisation of a hard copy of the donors’ results did not occur. Therefore it is possible that donors were non-compliant, or that the five samples in our study that were negative on all assays had a biological absence of antibodies.

All assays other than Euroimmun IgA displayed excellent specificity in testing a control cohort of pre-COVID-19 sera. The control cohort also included a small panel of non-SARS-CoV-2 sera positive for other respiratory virus antibodies, and we found no cross-reactivity on any of the assays used. Others have suggested that S1 and N are highly specific targets for SARS-CoV-2 serological analyses (23), and these antigens largely form the basis of current commercially available assays. A limitation of this work is that the timing of serum collection was not standardised, and that samples obtained were not tested equally across all assays due to limitations in sample volume and dead volume requirements of the automated EIAs. Nonetheless, the relatively large number of samples run remains a strength of this study, and allows for a head-to-head comparison of commercially available SARS-CoV-2 serological tests seen in few other studies.

We and others have shown that commercially available serological assays for SARS-CoV-2 have varying performance that is dependent on both the platform and marker used. The assay chosen by end-users should be tailored to specific applications - for example, assays measuring antibodies to the nucelocapsid antigen might be best suited to disease surveillence as spike-protein based vaccines become available. During the COVID-19 pandemic, serological assays will be crucial in answering questions of immune protection against reinfection. If convalescent plasma or COVID-19 immunoglobulin is found to be a potentially effective therapeutic intervention, high throughput serological assays that closely correlate with neutralisation antibody levels are vital for scalability. As further testing platforms become available, validation studies such as this are needed to identify assays that inform on antibody titre and functionality.

## Data Availability

Data related to this manuscript will be available to anyone immediately following peer-reviewed publication through contacting the corresponding author by email. These data include assay results for deidentified participant samples.

## Acknowledgements

The authors would like to thank the Clinical Chemistry Laboratory at the Prince of Wales Hospital Randwick, the Reproductive Endochrinology Laboratory at the Royal Hospital for Women Randwick, and University of New South Wales Stats Central for their generous technical support. Australian Governments fund Australian Red Cross Lifeblood for the provision of blood, blood products and services to the Australian community.

