## Supplementary Information for "SARS Coronavirus-2 microneutralisation and commercial serological assays correlated closely for some but not all enzyme immunoassays"

**This file includes:**

Supplementary method. SARS-CoV-2 spike RBD protein production

Table S1. Samples used to assess sensitivity and specificity of serological assays for SARS-CoV-2

**SARS-CoV-2 spike RBD protein production**

SARS-CoV-2 Spike RBD was cloned into pCAGGS as described previously (1). The plasmid was transiently transfected into Expi293-Freestyle cells (ThermoFisher Scientific) as follows: 1·5x10^8^ total cells (50mL transfection) were mixed with 50 µg of plasmid, 160 µL of ExpiFectinamine and 6 mL of OptiMEM-I and left overnight at 37°C in a shaking incubator. The following day 300 µL of ExpiFectamine Enhancer 1 and 3 mL of ExpiFectamine Enhancer 2 was added to the cells before the cells were left in culture for a further 48 hours. After a total of 72 hours in culture, the cell culture is collected and centrifuged for 20 minutes at 4000xg, 4°C. Cellular debris was clarified by passing the supernatant twice through a 0·22 µM filter. The His-tagged protein was then affinity purified from the cell supernatant using a HisTrap HP Column (GE Healthcare) and eluted with imidazole. The purified protein was then buffer exchanged and concentrated in sterile DPBS by centrifuging at 4000xg for 30 minutes at 4°C in a 10,000 MWCO Vivaspin centrifugal concentrator (Sartorius) and stored at – 80°C. The recombinant RBD was biotinylated using a Biotin Protein Labeling Kit (Roche). To prepare antigen-coated wells for the in-house ELISA, 8-well strips were initially coated with 100µL/well streptavidin (10µg/mL) and then blocked with 5% BSA in TBST. This was followed by coating with biotinylated RBD antigen (100µL/well, 10µg/mL in 0·5% BSA).

| **Table S1.** Samples used to assess sensitivity and specificity of serological assays for SARS-CoV-2 | | |
| --- | --- | --- |
| **Measurement** | **Confirmed Infection** | **Samples** |
| Sensitivity | SARS-coronavirus-2 | 200 |
| Specificity | N/A | 75 |
|  | Influenza virus A | 7 |
|  | Influenza virus B | 7 |
|  | Enterovirus | 5 |
|  | Respiratory Syncytial Virus | 3 |
|  | Adenovirus | 1 |
|  | Parainfluenza virus type 1 | 1 |
|  | Parainfluenza virus type 3 | 1 |
